# Expanding Hepatitis B Screening with Point-of-Care Rapid Testing in Primary Care: An Implementation Science Study

**DOI:** 10.1101/2024.08.29.24312788

**Authors:** Trang N. D. Pham, Long B. Hoang, Diem V. B Dao, Thao T. Dang, Van T. Nguyen, Duc H. Le, Thai N. Truong, Toan T. Le, Bao Q. Duong, Tram T. Trinh, Hang V. Dao, Doan Y Dao

## Abstract

**Background:** Vietnam faces a significant burden of hepatitis B virus (HBV) with around 10% of the population living with HBV and up to 80% unaware of their infection status. This study implemented a strategy using point-of-care rapid testing (POC-RT) for early HBV detection and linkage to care in primary care settings. The EPIS frameworks guided implementation, assessing barriers, enablers, feasibility, and acceptability.

**Method:** The implementation plan integrated insights from participating site’s authorities, local policies, practices, and patient pathways. A mixed-methods approach was employed at three primary care clinics in North Vietnam. Each site received 200-300 POC-RT test kits for use within 10 weeks. Patients received pre- and post-screening consultations for HBV using POC-RT, followed by referrals. Quantitative and qualitative data was collected to assess barriers, enablers, feasibility, and acceptability.

**Results:** Out of 600 POC-RT tests, 24 HBV positive cases were identified. Nine cases from public clinics received follow-up testing and specialist referrals within two weeks. Patients favored POC-RT for its simplicity, minimal blood requirement, and no cost. Physicians found POC-RT feasible for mass screenings, but noted challenges related to older adults’ skin thickness and blood volume accuracy. Linkage to care was satisfactory, but patients desired more health information during treatment.

**Conclusion:** Implementing POC-RT at primary care settings with comprehensive consultation and linkage to care is feasible and well-received. Patient feedback underscores the need for ongoing health information during treatment and follow-up, which is vital for retention in care. Enhancing the test kit design could further enhance POC-RT uptake in the population.

**Research in Context:** *Evidence before this study:* As of May 1, 2024, a comprehensive search on PubMed and Google Scholar using the terms (“HBV” or “viral hepatitis B” or “hepatitis B virus”) AND “point-of-care” AND “screening” AND “Vietnam” revealed no relevant studies specifically investigating the use of lateral capillary blood flow point-of-care rapid tests (POC-RT) for HBV screening in Vietnam. This highlights a significant gap in the literature regarding the implementation of POC-RT for HBV in this region.

*Added value of this study:* This study is pioneering in its application of the Exploration-Preparation-Implementation-Sustainment (EPIS) framework to incorporate POC-RT for HBV screening at the primary care level in Vietnam. It adopts a universal screening approach, recommending HBV screening for all patients irrespective of their risk profiles. This is a paradigm shift in HBV testing strategy in Vietnam. Until now, HBV testing is typically based on clinical signs and symptoms of HBV related end stage liver disease in Vietnam. As a result, only an estimated 20-30% of people living with HBV are diagnosed and thus 1.32% of the eligible are on anti-viral therapies. Our proposed study suggested shifting from a model of HBsAg testing based on clinical indication to a public health model of routine community wide HBsAg testing to achieve greater absolute and more equitable reach of HBV diagnosis across demographic characteristics. Additionally, the study documents how local authorities and healthcare workers have adapted to using POC-RT and the universal screening approach in both public and private primary care facilities at central and provincial levels. Furthermore, it identifies challenges related to test selection and patient linkage to care, while also considering patient perspectives on mass screening feasibility and acceptability.

*Implications of all the available evidence:* Given the high prevalence of HBV and the significant proportion of individuals unaware of their infection status and unscreened for HBV, Vietnam would benefit from a proactive universal screening strategy. Decentralizing screening services using POC-RT at the primary care level is strategically essential toward this universal premise. Future research should focus on broader implementation and dissemination of POC-RT in primary care and other resource-limited settings, with an emphasis on effective methods for ensuring linkage to care. Taken together, this study’s novelty lies in its systematic approach to integrating POC-RT in Vietnam’s healthcare infrastructure, potentially setting a precedent for similar interventions in other regions with high HBV prevalence and limited resources

## Introduction

Chronic hepatitis B virus (HBV) infection remains a significant global public health challenge, with approximately 290 million individuals living with HBV worldwide.^1,2^ Without intervention, patients with HBV have an approximately 30-40% lifetime risk of developing liver cirrhosis or hepatocellular carcinoma (HCC).^3-5^ Vietnam, a low- or middle-income country in the Asia Pacific with a population of 97 million, bears one of the largest burdens of HBV, with 7-10% of the Vietnamese population living with HBV.^6,7^ In 2022, only 30% of adults in Ho Chi Minh City (HCMC), a city with 10 million persons, were tested for HBV.^8^ Of the 30% who were tested for HBV, 75% were diagnosed incidentally (i.e., hospitalized for other medical conditions (40%) and liver cirrhosis decompensation or advanced liver cancer (60%).^8^ Nationally, only 13% of individuals estimated to be living with HBV were aware of their infection.^9^

HBV testing and care access is one of five core interventions to eliminate the global viral hepatitis threat by 2030, which are impactful, affordable, and cost-effective. The other four measures related to prevention (childhood vaccination, prevention of mother-to-child transmission of HBV, blood and injection safety, and comprehensive harm reduction for people who inject drugs) have been integrated into Vietnam’s immunization program and HIV/AIDS programs over the past 20 years.^10^ For HCC prevention, diagnosis, and treatment of HBV is well-documented as one of the most effective strategies. However, more than 98% of the eligible HBV treatment recipients are untreated in Vietnam.^11^ One reason for the low treatment rate is the lack of available and accessible testing in primary care settings. Currently, most HBV testing in Vietnam is phlebotomy-based, and follow-up management (viral load testing, liver function and fibrosis assessment, care management, etc.) is concentrated at tertiary or secondary care levels. The provision of HBV testing and diagnosis services in primary care at district-level outpatient clinics is negligible and underutilized by patients, partly due to the lack of formal recommendations from primary care providers.^8^ These factors contribute to the low HBV testing and treatment rates.

Scaling up testing and improving linkage to care will require developing and implementing innovative strategies to overcome systemic barriers to hepatitis B diagnosis and treatment access.^12,13^ Point-of-care rapid testing (POC-RT) for HBV is key to increasing screening and accessibility to care, especially in primary care facilities. POC-RT is a specimen collection method with advantages including simplicity and relative ease of collection by finger prick, preparation, transport, and storage.^12^ These tests are indirect immunoassays with a sensitivity and specificity similar to FDA-approved laboratory-based HBV antibody assays. With its fast turnaround time (about 20 minutes), POC-RT enables timely linkage of patients to further assessment within one contact with the health service, reducing the lost-to-follow-up to viral load testing and treatment initiation.^12,13^. A feasible and potentially effective strategy is to provide access to testing through POC-RT for HBV in primary care clinics – the first point of entry into the healthcare system – in Vietnam.

Taken together, we conducted an implementation study to test the feasibility and acceptability of decentralizing POC-RT HBsAg testing and subsequent care access for HBV in district-level clinics in Vietnam. The results of this study will inform the design and development of a sustainable scale-up implementation project. We applied the implementation science’s EPIS (Exploration – Preparation – Implementation – Sustainment) framework^14^ to explore the barriers and enablers of implementing POC-RT in selected primary care settings and the project’s reach, adoption, and implementation. This report focuses primarily on the implementation phases of the project.

## Methods

### Study design and setting

Between July and November 2022, we conducted implementation research using a mixed methods approach at three primary care facilities in Vietnam. The sites included two primary care clinics within public hospitals—Dong Da General Hospital (DDH) in Hanoi and Thanh Hoa City General Hospital (THH) in Thanh Hoa Province – as well as one private gastroenterology and hepatology clinic–Hoang Long Clinic (HLC) in Hanoi (Figure 1). These sites were intentionally selected to represent typical accesses points for primary care services across both public and private healthcare sectors at urban and provincial levels, their capabilities to provide hepatology services as referrals for patients after HBV screening. Detailed characteristics of the three sites are presented in Table 1.

**Table 1.**
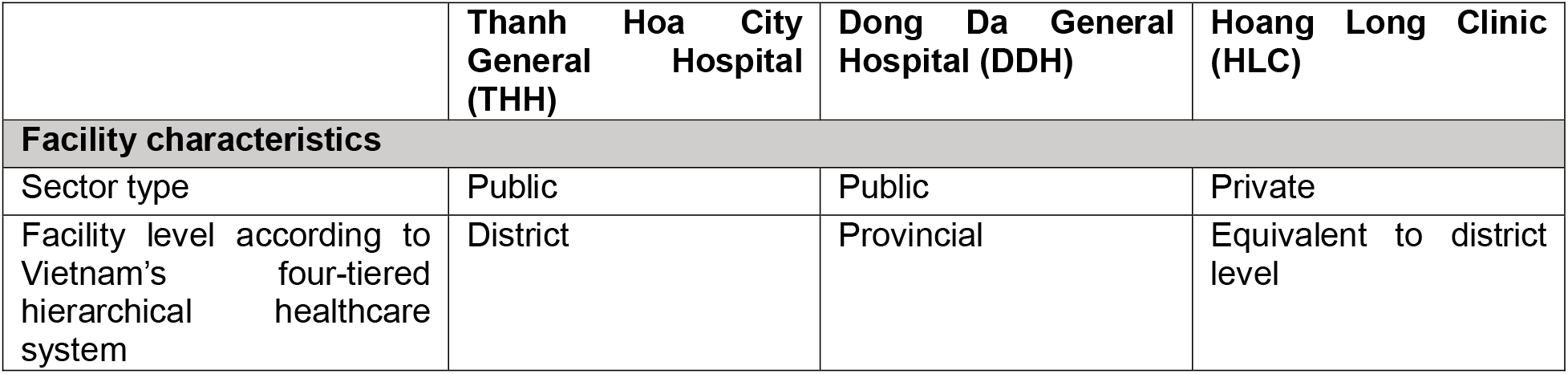
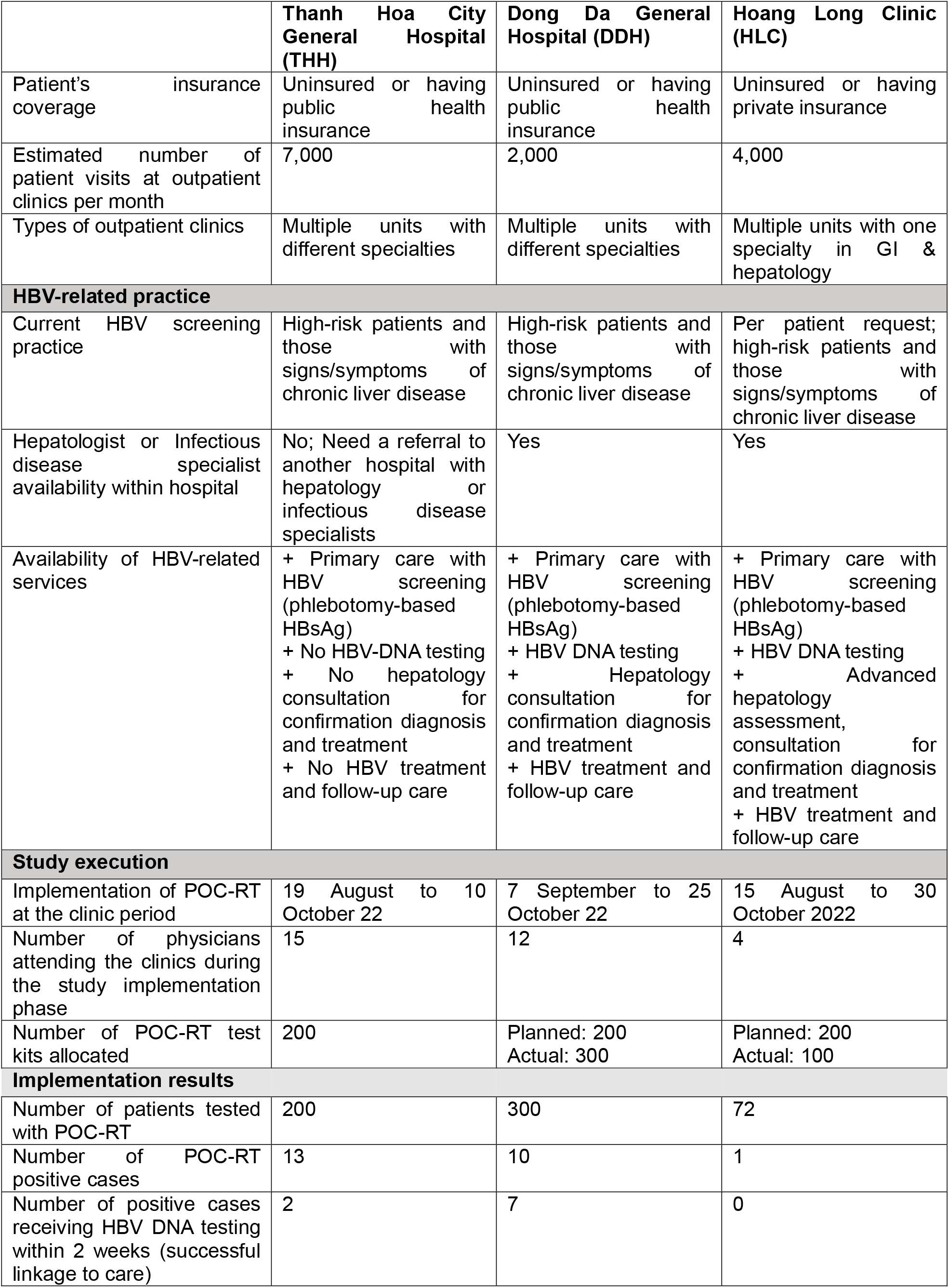
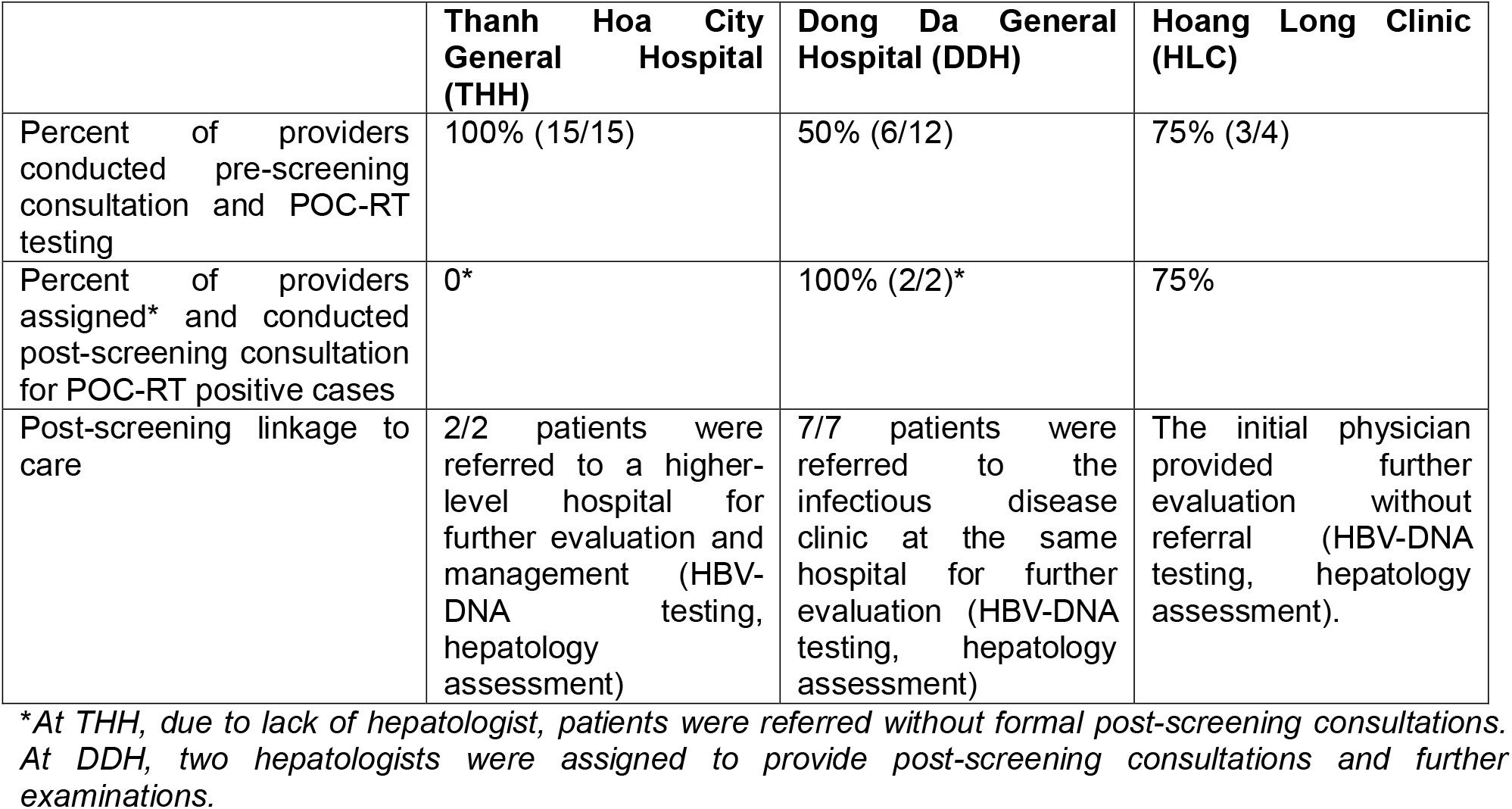
Participating sites characteristics and services outcomes.

**Figure 1.**
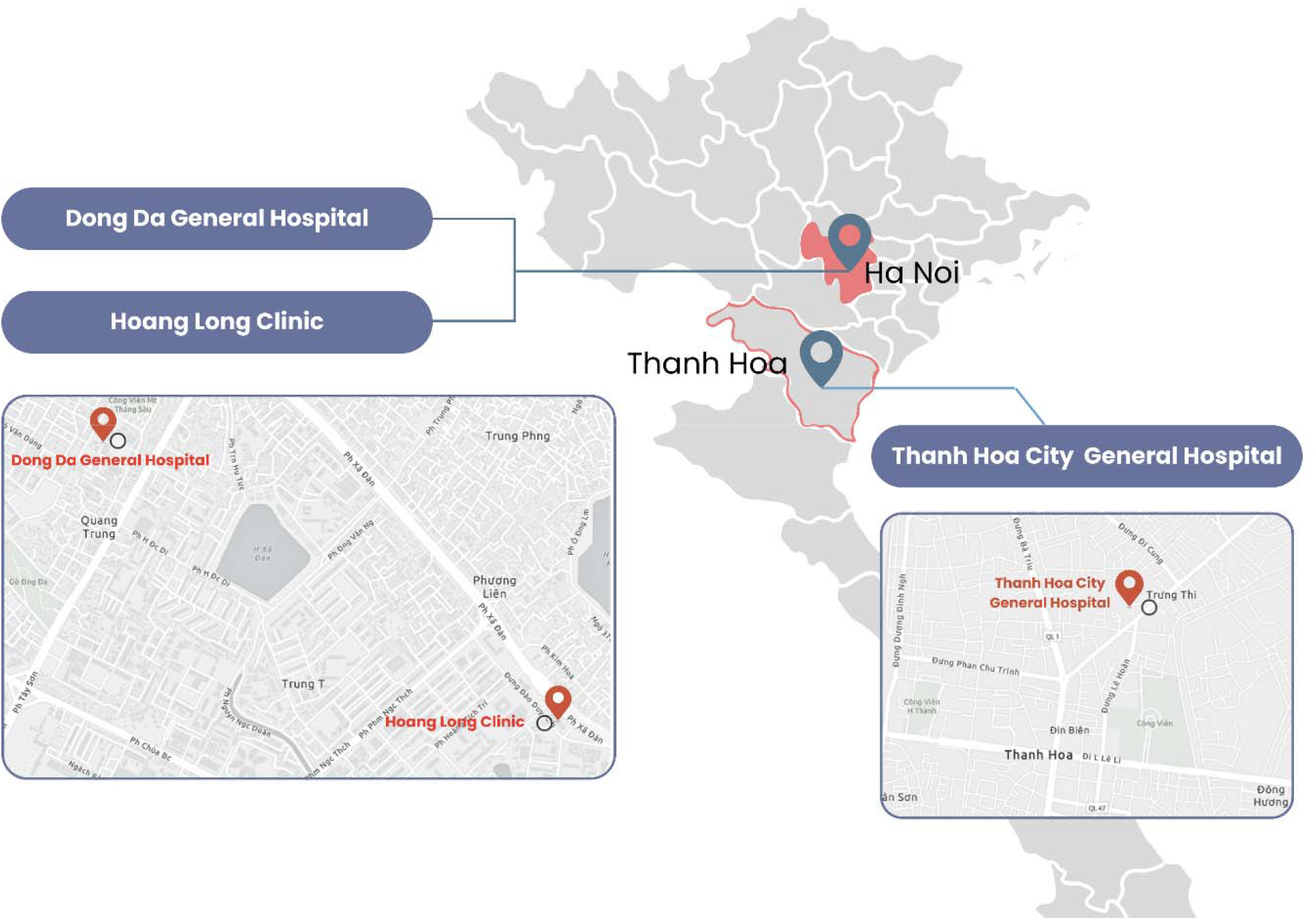
Locations of the primary healthcare factilities study sites in Vietnam.

The study protocol was approved by an institutional review board (IRB)—Dinh Tien Hoang Institute of Medicine in Hanoi, Vietnam (approval number IRB-2201 dated 03/05/2022).

### Study population and patient identification

Our study targeted both patients and healthcare providers. Patients seeking care at the clinics, aged 18 or older, were eligible to participate. Consent was obtained before HBV screening and for subsequent in-depth interviews (IDI). Patients who had a mental illness or a severe comorbidity and had a history of HBV vaccination were excluded. Healthcare providers who participated in the study activities were included in focus group discussions (FGD). Providers and patient characteristics were reported in Table 1.

### Study implementation and data collection

#### Exploration and Preparation

One month before finalizing the study’s operational procedures, we conducted a pre-study field assessment. Guided by the EPIS framework, we adapted operational procedures based on findings from our prior exploration.^14^ These findings included leadership priorities and support, facility awareness, care provider characteristics and willingness to change, and the patient journey and clinical pathway (Appendix A).

As per the agreement with the IRBs and site directors, each participating facility integrated POC-RT into their usual practice. They provided a fixed number of POC-RTs (10 tests/day) free of charge for participants, along with pre- and post-screening consultations. By limiting the number of free tests per day, we ensured that care providers could adhere to the protocol without overwhelming the current workflow at the sites.

The POC-RT HBV test used in this study was SD Bioline HBsAg WB^®^, manufactured by Abbott Rapid Diagnostic. This test received WHO’s prequalification of In Vitro Diagnostics.^15,16^ A study assessing diagnostic accuracy showed that they had high sensitivity (98·1% to 100%) and specificity (97·2% to 99·8%).^17^ Vietnam’s Ministry of Health (VN-MOH) approved the test for clinical use in 2017.^18^ We procured the POC-RT before initiating patient recruitment and provided testing instructions and quality assurance information to all participating sites.

Local hepatologists, experienced in current practices and patient cultures, prepared comprehensive pre- and post-screening consultation information for all participating sites. The pre-screening consultation covered the high prevalence of HBV in Vietnam, clinical benefits of screening and treatment, who should be tested, advantages of POC-RT over lab-based tests, test procedures, and what patients can expect if they test positive. The post-screening consultation included result explanation, the need for confirmed diagnosis or vaccination, encouragement to screen family members, potential follow-up costs, treatment initiation, and public health insurance details.

#### Implementation

Each participating site was allocated 200 POC-RT test kits for use in their respective outpatient clinics. All physicians and nurses at these sites underwent training in the study protocol, including in-office POC-RT procedures, quality assurance, and pre- and post-screening consultation. Nurses also received additional training for patient care linkage and progress documentation. Each site independently established its POC-RT implementation timeline between September and November 2022.

Physicians introduced patients to HBV screening during pre-screening consultation. Those who agreed to use POC-RT (rather than lab-based testing) were included in the study. After obtaining consent, patients used POC-RT to screen for HBV under nurse supervision. Patients then returned to their initial physician for evaluation and post-screening consultation. If the screening results for HBV were negative, patients were provided with information on HBV infection prevention, including vaccination. For positive results, care linkage was established, referring patients to within-hospital hepatologists, infectious disease specialists, or external hepatologist or lab for confirmation diagnosis and further management. Patients who received care linkage recommendations were followed up for two weeks to assess the success of patient linkage. If patients did not undergo follow-up measurements (viral load testing, hepatology assessment, and consultation with an infectious disease specialist or hepatologist) within two weeks after screening, we contacted them again after an additional two weeks to confirm the success of linkage. Testing and linkage-to-care data were documented by participating site nurses.

IDIs were conducted with patients who tested positive using the POC-RT. Each IDI lasted between 30 and 45 minutes. Focus group discussions were conducted among healthcare workers who participated in the study. We conducted one FGD for each site, and this discussion lasted about 1 hour. Both the IDIs and FGDs were based on a semi-structured questionnaire, exploring participants’ experiences with the POC-RT HBV test, linkage to care, and their viewpoints on scaling up POC-RT testing among patients and healthcare workers.

#### Sustainment

Information from qualitative interviews with patients and care providers was extracted and combined with end-of-project research team meetings to gain knowledge about improvement for scaling up for the upcoming implementation project and measures to sustain the implementation of POC-RT at the primary care level.

### Sample size estimation

We aimed to screen 600 patients using POC-RT for quantitative data collection, anticipating approximately 48 positive cases for care linkage. This estimation was based on an 8% HBV prevalence in the population,^8^ with a standard error of 6% at a 95% confidence interval, and an estimated 73·9% of patients continuing their care at the referral destination.^19^

For qualitative data collection, we conducted one FGD at each participating site, involving 6-8 care providers (physicians and nurses) per site. Patients aged 18 and older who tested positive for HBV using POC-RT were invited for IDIs. We ensured patient representation across genders, age groups (18-40, 41-60, and 61 or older), and education levels (below high school graduate, and at or above high school levels).

### Outcomes assessment and data analysis

The project’s outcomes was assessed based on the provision of services, which was documented and calculated as follows (see Table 1, implementation results): (1) the number of patients tested with POC-RT, (2) the number of POC-RT positive cases, (3) the number of positive cases that underwent HBV-DNA testing within two weeks (defined as successful linkage – the HBV-DNA test was done within two weeks post screening), (4) the percentage of providers who provided pre-screening consultation and POC-RT, (5) the percentage of providers who provided post-screening consultation for POC-RT positive cases, and (6) the measure of post-screening linkage to care. Documented data was obtained from the hospital’s health information system and verified with the site coordinator’s progress tracking note. The quantitative data was reported as documented for descriptive purposes. No hypothesis statistical testing was conducted.

Variations in implementation (clinical pathway), technical issues (testing procedure, the necessity of professional personnel to perform and interpret the test, potential of large-scale use), acceptability, linkage to care, and potential for scaling up were derived from qualitative data. All IDIs and FGDs were audio-recorded. Any identifiable patient information was coded before the recorded discussions were transcribed verbatim, coded and analyzed using the deductive-inductive method. The thematic approach was used to draw contextual information and details about the implementation process. Microsoft Word and Excel were utilized for qualitative data organizing, coding, and analysis. Codes and themes were cross-checked.

## Results

### Implementation of HBV POC-RT

We implemented the HBV POC-RT at three study sites with diverse profiles in sector types (both public and private), healthcare level (district and provincial level), insurance coverage (with and without public health insurance coverage), and expertise and facilities for hepatology and infectious disease evaluation. (Table 1)

All sites provided HBV screening using HBsAg, mostly for high-risk patients and/or those having symptoms and signs of chronic liver disease. Basic testing for hepatology evaluation (transaminase enzymes, bilirubin) was available in all sites, but HBV-DNA quantification was not available at THH, thus, HBV-positive patients had to be referred to a higher-level hospital (provincial level hospital) for HBV-DNA testing, further diagnosis, and HBV management. Although all physicians received training in pre- and post-screening consultation and care referral, each hospital designated specific providers for post-screening consultations. Initially, we planned to provide each site with 200 POC-RT test kits; however, owing to slow recruitment at HLC, we later transfered 100 test kits at HLC to DDH. A total of 572/600 (95·3%) test kits were used.

#### Acceptability

After receiving pre-screening consultation, all patients at the two public hospitals agreed to use POC-RT rather than phlebotomy-based tests; consequently, all allocated tests were completed within the allotted time, However, at the private clinic (HLC), the majority of patients with known HBV status and physicians preferred phlebotomy-based tests over the POC-RT.

Patients reported that using POC-RT with only three finger-pricked blood drops was less frightening than using 1-2 venous blood samples for phlebotomy-based testing. In FGDs, care providers stated that the POC-RT was <u>adaptable</u> to their practice as the clinical pathway for HBV screening using POC-RT was similar to their routine phlebotomy-based test procedures, which included taking a history of HBV vaccination and family history and performing a physical examination. The main difference was that they provided more information about HBV POC-RT during the pre-screening consultation, emphasizing it as a valid alternative to phlebotomy-based tests. They also highlighted several advantages of POC-RT, including requiring only a fingerstick instead of phlebotomy, making it more patient-friendly. At times, care providers mentioned the free-of-charge testing under the current research program as a motivation for patients. They often promoted *“the test is completely free”* and spent more time explaining the program to patients than usual.

The patient’s perspective on pre-screening consultation and POC-RT was mainly focused on “accuracy, convenience, rapidity, free, and fingerpick”. Care providers stated that the simple testing procedure and “*available right away*” of test results were the main factors contributing to the <u>feasibility</u> of using POC-RT: “*…it [POC-RT] saves time and a lot of human resources. Per the old [current routine] protocol…the process is very lengthy, involves many steps, and requires a lot of human resources*” (a care provider at a public hospital). However, some providers did not consider POC-RT to be a replacement but rather as an alternative to phlebotomy-based test and expressed uncertainty about the POC-RT’s diagnostic accuracy concern. In addition, at HLC where the phlebotomy-based HBV test is available and often prescribed together with hepatitis C and other viral tests as a service package, the physicians were discouraged to separate the package to prescribe the POC-RT for HBV testing. Most care providers agreed that POC-RT could be used in mass screening for hepatitis B (see detailed in Possibility to scale up) or when no other tests were required.

All three healthcare institutions reported <u>technical difficulties</u> when they performed POC-RT with various types of lancets, especially in elderly patients who often have thicker skin to puncture. Lancets with narrow and thin needles were not appropriate to use due to their shallow penetration, taking a long time to get adequate blood for testing (“*but the drops are small, it will clot immediately at the well…because this blood is not anticoagulated…”)*. A second concern with POC-RT was the test well was small and deep, making it difficult to exactly drop the blood at the center of the well and fill it up (“…*If there was not enough blood, the test would not run. The well must be full [of blood]*”).

Care providers and patients agreed that POC-RT <u>can be scaled up</u> for mass testing due to its simplicity, low cost, and quick turnaround time, especially in rural and low-resource settings. To facilitate the uptake of the test, healthcare workers emphasized the importance of patient education about the risks of HBV and the importance of HBV early detection. Promotional messages should include the following phrases: “hepatitis B is an infectious disease”, “hepatitis B results in severe health consequences”, and “screening for HBV helps to avoid transmission to other people”.

Regarding <u>sustainability</u>, care providers shared some insights about the challenges in implementing POC-RT at the hospital’s outpatient clinics. First, the overcrowding status at some hospitals limited the time for thorough consultation and prolonged the waiting time for the POC-RT test to be done individually at the laboratory. Nurse-led office-based POC-RT should be considered for future implementation. Second, post-screening consultation should focus more on result interpretation and risk communication. To do so, consultation time should be secured and paid, ideally under public insurance coverage and national guidelines. Third, developing more user-friendly designs for the POC-RT kit would facilitate self-testing at home (like COVID-19 test). In addition, instructions for the testing procedure should be delivered in multiple forms (i.e., a small brochure, or a video illustration). Fourth, with the increasing network of pharmacies nationwide, training pharmacist assistants to consult and perform POC-RT would be a potential strategy to increase HBV screening uptake. Finally, to encourage POC-RT use at hospital clinics (similar to the use of pregnancy rapid test), scientific evidence with proof of local verification should be provided to physicians to ascertain the diagnosis accuracy of POC-RT and accept its use in replacement of phlebotomy-based test.

### Linkage to care – service uptake, adaptation, and patient satisfaction

All HBV-positive patients (n=24) received post-screening consultations regarding linkage to care from their primary care providers. Of these, nine patients received further evaluation (HBV-DNA testing and hepatology assessment) and started treatment within nine months after testing (7 at DDH and two at THH). The only patient who tested positive at HLC refused to perform further evaluation due to the insurance reimbursement restrictions. Among those who tested positive but did not complete follow-up testing within two weeks after screening, the reasons for not doing so included (1) being busy with family and work, (2) feeling that further evaluation was not urgent because other test results were normal, (3) not being ready for treatment despite being aware of the disease, and (4) financial considerations. Finance was an important theme, repeatedly discussed in the FGDs—for example, at the hospital where THH patients were referred to, the HBV-DNA quantification costs US$ 72 (for reference, per capita in Vietnam is US$ 336)^20^ and would not be covered by health insurance for the first evaluation after the detection of positive HBsAg. At many private clinics, patients were not covered by public health insurance and would have to pay out-of-pocket for all the further examination tests (“…*If we do all the tests for patients, such liver fibrosis, viral load, other serum markers, and vitamin D and so on, it will cost a few hundred US dollars. For financially competent patients, it’s okay, but there are many patients who can’t afford that amount”*).

There were slight <u>variations in adaptation</u> of care linkage across participating sites. While most of physicians provided pre-screening consultation (half of the physicians in DDH did not). The post-screening linkage to care differed among the three sites because of some factors. At DDH, where infectious disease specialists were available within the hospital, positive cases were referred to the outpatient clinic of the Infectious Disease Department, and they received further testing (such as viral load and liver fibrosis) and consultation. At HLC, GI and Hepatology providers consulted positive patients about the need to perform additional testing and treatment. At THH, patients were referred to outside higher-level hospitals for further testing and treatment.

Some doctors reported that they also provided psychological support for patients who worried about their positive HBsAg status so that they were motivated to perform additional testing to start treatment. They discussed the risks of transmission and liver cancer development and advised that patients’ family members should also be tested.

Although HBV <u>patients were generally satisfied</u> with the linkage-to-care procedure, they also expressed the need to know more information about treatment and management after the confirmation of HBV-DNA (which was not included in the post-screening consultation). Particularly, they wanted to know more about the centers that treat HBV in their area, treatment, and medication options, how treatment will be decided based on their test results, modes of transmission, risks to themselves and family members, how to prevent infection and transmission, and whether the disease can be cured. One patient voiced the frustration caused by overcrowded clinics: “*…The hospital is too crowded, I’m uncomfortable, and so is the doctor…I want to know more about the medications that I use. They just give me a prescription, tell me to buy the medications, I only know [that I have to take] the medications but don’t know what they are and how to use them…I have to read the package insert [to find more information about the medications]. We don’t know medical terms, we don’t know drug names, while the prescription is so long and full of drug names that we don’t understand”*). Thus, it was suggested that information regarding treatment and management for confirmed HBV-positive patients should be focused on retaining patients in care and improving patient care adherence in scaling up.

## Discussion

Our study showed that it is feasible to implement HBV screening with POC-RT at primary care-level outpatient clinics. Applying the EPIS framework helped us gain first-hand experience and insights into the local settings and engagement of facilities leadership and healthcare providers, making the study protocol more easily adapted to daily practice. Patients and providers accepted the use of POC-RT for HBV screening. Post screening with POC-RT care linkage within two weeks of testing illustrated a potential strategy to retain patients within care, especially at hospitals outpatient departments with attending hepatologists or infectious disease specialists.

HBV screening with POC-RT can be leveraged to overcome challenges in testing accessibility (cost, availability, simplicity, ease of use, quick turnaround time) within Vietnam’s universal healthcare system, where the primary care system forms the backbone. POC-RT should be implemented at district-level primary care clinics because primary care physicians provide first-contact care and gatekeepers to health and social care systems. Integration of POC-RT for HBV and access to HBV treatment is within routine primary care service delivery and policy support, reimbursement, infrastructure, and tools that are already in place. Furthermore, over 90% of the Vietnamese population have health insurance, which covers 80% of treatment costs, indicating that HBV-related end-stage liver disease can be prevented by delivering HBV testing and treatment to primary care levels in Vietnam.^21^ The implementation of hypertension and diabetic screening at the primary care level in the community path the way for the scalability of decentralized HBV screening with POC-RT in Vietnam.^22,23^ In addition, using POC-RT at pharmacies is a potential strategy to consider in future implementation plans. The availability of HBV POC-RT in the market, with Vietnam’s MOH approval, makes it more feasible for hospitals to procure these tests.

At the time of writing, HBV screening is not reimbursed by public health insurance in Vietnam. The main goal of advocating the integration of HBV POC-RT and access to HBV treatment into routine primary care service delivery is to enhance the early detection and treatment of HBV. With the low cost of POC-RT (fourth compared to lab-based screening tests), subsidized or sliding fee scale screening coverage from public insurance may increase testing uptake in the community due to its ease of use and quick turnaround time. In addition, including these services in the public health insurance scheme ensures they are accessible and affordable for the general population. This approach can lead to increased screening rates, better linkage to care, and improved treatment adherence, ultimately contributing to the reduction of HBV transmission and the incidence of liver cancer. The availability of these interventions within primary care also supports the goal of eliminating HBV as a public health threat, in line with global health goals.

Implementing POC-RT is not enough to increase HBV screening uptake. Knowledge about HBV infection and prevention is modest in the general population, emphasizing the need for public health promotion and community health education.^8^ Further, the current practice of prescribing HBV screening tests does not involve formal pre- and post-screening consultation.^24^ In addition, care providers are lack of awareness of the availability and accuracy of POC-RT in HBV screening, opening opportunity to endorse for use in replacement of phlebotomy-based tests by the Vietnam’s Ministry of Health and WHO. Lessons from HIV prevention efforts underlined the significant changes in screening behaviors with pre- and post-screening consultation combined with HIV health education and promotion.^25^ These strategies should be considered in implementation efforts to improve HBV screening. In addition, testing consultation, result interpretation, and risk communication enhance service delivery, build trust, and ensure patients receive sufficient in-care services.

Post-screening linkage to care was well-received, but the demand for health information related to HBV treatment and management arose. Patient’s need for treatment and follow up information were not met during visits due to limited time for consultation. HBV treatment requires long term therapy to suppress HBV replication effectively.^26,27^ Thus, poor adherence to treatment was associated with an increased risk of virologic relapse.^26,27^ Studies indicate that 40-60% of patients could not accurately recall their physicians’ instructions within 10-80 minutes of receiving the information.^28^ Further, more than 60% of patients misunderstood the directions for prescribed medication immediately after their doctor visits.^29^ Thus, with the limited time for consultation during health visits, poor patient-provider communication could contribute to non-adherence.

### Strength and limitations

By applying the EPIS framework, we gained insights into the local settings and practice, selected approaches to engage stakeholders, and better prepared for the implementation phases. Our protocol was quickly adapted to similar settings due to the hospital authorities’ support and approval, well penetration of the study protocol to care providers, and the study operational procedures were similar and easily integrated into routine clinical pathways. This study demonstrated that it is necessary to have guidance from an implementation science framework to address multilevel factors contributing to the success of a project. However, generalizing our findings to outside the participating sites should be approached with caution.

## Conclusion

In conclusion, the pilot study has laid the groundwork for further research and implementation projects aimed at improving HBV testing accessibility and linkage to care in Vietnam. The EPIS framework provided valuable guidance in leveraging innovative strategies and addressing systemic barriers in real-world implementation. POC-RT for HBV screening can be integrated into routine primary care service delivery, especially at district-level primary care clinics. The availability of HBV POC-RT in the market, with approval from VN-MOH, makes it feasible for hospitals to procure these tests. This study also underscored the need for public health promotion and community health education to increase knowledge about HBV infection and prevention. Additionally, formal pre- and post-screening consultation, along with improved patient-provider communication, are essential to enhance service delivery and ensure patient adherence to treatment after getting screened. It is necessary to advocate for the integration of HBV screening and treatment into routine primary care service delivery, along with potential reimbursement for screening from public health insurance, can contribute to increased testing rates and better linkage to care. By addressing multilevel factors contributing to the success of the project, it is possible to make significant progress towards the goal of eliminating the global viral hepatitis threat by 2030.

## Data Availability

All data produced in the present study are available upon reasonable request to the authors

## Acknowledgments

Johns Hopkins GI and Hepatology Division’s Pilot Fund

Johns Hopkins’ Center of Excellence for Liver Disease in Vietnam

Mai Dolch Foundation

