## Appendix for "Expanding Hepatitis B Screening with Point-of-Care Rapid Testing in Primary Care: An Implementation Science Study"

**Appendix A**. EPIS framework, sources of information gathered during the Exploration phase and activities in Preparation and Implementation phases, and subsequently lessons learned for Sustainment.

**Exploration**

**(Field assessment):**

+ Leadership engagement

+ Facility characteristics

+ Procedure

+ Patient journey

+ Clinical pathway

**Preparation:**

+ Approval from hospital IRB & board of director

+ Consensus on clinical pathway and patient journey for the study period

+ Secure test kits for implementation period

+ Obtain agreement on caseload and services provided

+ Training providers in study procedures, pathway and consultation.

**Implementation:**

+ Each hospital IRB approved the protocol and clinical pathway and POC-RT testing load.

+ Allocation of test kits

+ Monitoring and recording progress

+ Evaluation (conduct in-depth interviews or focus-group discussions)

*Lessons learned for Sustainment*
